# Medication adherence and functional food use in patients with hypertension: a cross sectional study

**DOI:** 10.1101/2025.08.20.25334126

**Authors:** Hidehiro Someko, Takeo Nakayama, Shiho Koizumi, Carl B Becker, Takahiro Tabuchi, Shuhei Ishikawa, Yousuke Yamamoto

**Author notes:** Corresponding author: Hidehiro Someko, Science and Technology in Public Sphere, Graduate School of Medicine and Public Health, Kyoto University, Yoshida Konoe-cho, Sakyo-ku, Kyoto 606-8501, Japan.

## Abstract

**Background:** Functional foods are increasingly used by patients with chronic diseases, including hypertension. However, the relationship between functional food use with therapeutic intent and medication adherence remains unclear, particularly among hypertensive patients.

**Methods:** This cross-sectional study analyzed data from the Japan COVID-19 and Society Internet Survey (JACSIS2024) conducted between December 2024 and January 2025. Participants were included if they reported having hypertension and were currently receiving medical care with medication. Those with ineligible responses were excluded. Participants were classified into functional food users and non-users based on their use of functional foods specifically for treating hypertension. Medication adherence was assessed using the Japanese version of the 8-item Morisky Medication Adherence Scale (MMAS-8). We estimated risk ratios (RRs) for low adherence (MMAS-8 < 6) associated with functional food use using modified Poisson regression with a log link and robust standard errors, adjusting for demographic, socioeconomic, and health-related covariates. Analyses were weighted to adjust for potential selection bias in the Internet-based sample.

**Results:** A total of 4,063 hypertensive patients receiving medication treatment were analyzed, comprising 586 functional food users and 3,477 non-users. In adjusted and weighted analyses, functional food use was associated with a higher risk of low adherence: RR 1.24 (95% CI, 1.04–1.48).

**Conclusions:** Hypertensive patients who use functional foods with therapeutic intent demonstrated poorer medication adherence compared to non-users. These findings suggest that functional food use with therapeutic intent may be associated with suboptimal medication adherence, potentially reflecting underlying patient attitudes toward conventional medical treatment. Further longitudinal research is needed to establish causality and explore whether targeted educational interventions could help optimize both functional food use and medication adherence in hypertensive patients.

## Introduction

Hypertension remains one of the most prevalent chronic diseases worldwide, affecting an estimated 1.28 billion adults globally^1^. As a major risk factor for cardiovascular diseases, stroke, kidney failure, and premature mortality, hypertension requires consistent, evidence-based medical management to achieve adequate blood pressure control. Despite significant advances in pharmacological treatments with proven efficacy in reducing morbidity and mortality, hypertension control rates remain suboptimal across most populations. Effective management necessitates not only appropriate medication prescription but also patient adherence to treatment regimens, as inconsistent medication use significantly diminishes therapeutic benefits and increases the risk of serious complications such as myocardial infarction and cerebrovascular events^2^.

Functional foods are broadly defined as food products that provide health benefits beyond basic nutritional content^3^. While this concept might appear modern, recognition of foods with specific health-promoting properties dates back centuries in various traditional medicine systems worldwide ^4^. The formal categorization and commercialization of functional foods emerged only in the early 1990s, subsequently spreading to wealthy consumer markets across Europe, North America, and parts of Asia. The sector demonstrates substantial economic significance, with the global functional food ingredients market valued at USD 128.12 billion in 2025 and projected to reach approximately USD 201.49 billion by 2034, growing at a compound annual rate of 5.19%^5^. Consumers are increasingly drawn to these products as part of their health management strategies, particularly for preventive purposes including cardiovascular wellness. Many perceive functional foods as natural complements to conventional health approaches, though their optimal role is as supplements to—rather than substitutes for—evidence-based medical interventions when treating diagnosed conditions like hypertension.

Despite the legitimate preventive benefits of functional foods, a concerning trend has emerged where patients with diagnosed conditions like hypertension use these products with therapeutic intent rather than as complementary approaches. Research indicates that 7.7-28.2% of patients use dietary supplements to treat their conditions, with many using them concurrently with prescribed medications^6^. A critical concern is that patients may believe functional foods can substitute for antihypertensive therapy or enable them to decrease prescribed medication dosages while avoiding pharmaceutical side effects.

The relationship between functional food use and medication adherence remains complex and not fully understood. While concerns exist that patients using functional foods may demonstrate reduced adherence to prescribed medications, this relationship has not been definitively established in the literature ^7,8^. Limited research exists examining how patients’ therapeutic use of functional foods might impact their adherence to prescribed antihypertensive medications, particularly when patients view these products as treatment alternatives rather than preventive measures. Studies focusing specifically on hypertensive patients are particularly scarce, creating a significant knowledge gap in this population.

Given this knowledge gap, the present study aimed to investigate the relationship between functional food use with therapeutic intent and medication adherence among hypertensive patients.

## Methods

### Study design and settings

This study utilized a cross-sectional design using data from the Japan COVID-19 and Society Internet Survey (JACSIS), a large-scale longitudinal internet-based cohort study established in August 2020 to investigate the multifaceted impacts of the COVID-19 pandemic on Japanese society. The present analysis used data from Wave 9 of JACSIS (JACSIS2024), conducted between December 2, 2024, and January 16, 2025. This study is approved by ethics committee of Kyoto University Graduate School of Medicine (R4810).

Participants were recruited through a two-step process managed by the commercial research agency Rakuten Insight, whose panel comprises approximately 2.2 million individuals from the Japanese population. First, all individuals who had participated in any previous wave of JACSIS (2020-2023) or the Japan “Society and New Tobacco” Internet Survey (JASTIS, 2015-2024) were invited to rejoin the study. Details of the JASTIS study design have been described elsewhere^9^. Among 45,359 contactable previous participants, 26,097 responded (response rate: 57.5%). To reach the target sample size of 28,000, an additional 1,903 new participants were recruited from Rakuten Insight’s panel using stratified sampling by sex, age, and geographic region. The online survey was administered via Rakuten Insight’s secure web platform, with participants receiving compensation for their participation.

### Regulatory framework of functional foods in Japan

Japan’s functional foods regulatory framework encompasses three main categories with distinct approval processes and purposes^10^. Foods for Specified Health Uses (FOSHU) are products containing ingredients with verified claims of physiological effects on the human body, requiring government evaluation and approval from the Secretary General of the Consumer Affairs Agency for their effects and safety before marketing. FOSHU products are available for health maintenance and promotion, or for consumption by those who wish to control a health condition. Foods with Function Claims (FFC) operate under the food industry’s own responsibility, applying to foods with structure and function claims based on scientific evidence, but unlike FOSHU, these products do not require individual pre-approval from government authorities. Foods with Nutrient Function Claims (FNFC) are used to supplement or complement the daily diet to ensure that nutritional requirements are met, covering 13 vitamins, 6 minerals, and n-3 fatty acids with well-established functions that can receive nutrient function claims without government notification. Despite this regulated framework, many functional foods exist outside these categories, and Japanese consumers often refer to all these products collectively as “supplements,” though this term has no legal definition in Japan and is generally accepted as equivalent to “health food.”

### Participants and Eligibility Criteria

From the initial 28,000 respondents of JACSIS2024, participants with ineligible responses were systematically excluded based on predefined criteria to ensure data quality. Ineligible responses were defined as meeting any of the following conditions: (1) survey completion time under 600 seconds (10 minutes), indicating insufficient attention to survey content (n=186); (2) incorrect response to an attention check question asking participants to “select the second-to-last option” (n=2,655, including 2,571 who selected the second option and 84 who selected other incorrect options); (3) reporting current presence of an implausibly comprehensive set of medical conditions simultaneously (11 out of 20 specific conditions, n=126); or (4) reporting household size exceeding 15 people, which exceeds realistic population distributions (n=6). After excluding 2,918 participants meeting these criteria, 25,082 participants remained eligible.

For the present study, participants were further restricted to those who reported having hypertension and taking medication based on their response to the survey item “Do you currently have hypertension?” as “Currently have it - receiving medical care with medication.”

### Data Collection and Measurements

The primary outcome of this study was medication adherence, which was assessed using the Japanese version of the 8-item Morisky Medication Adherence Scale (MMAS-8)^11^. The MMAS-8 is a validated self-report instrument designed to identify barriers to and behaviors associated with medication adherence. The Japanese version has demonstrated acceptable reliability and validity, with a Cronbach’s α of 0.74 in patients with ulcerative colitis and good convergent validity with self-reported missed doses (Spearman’s correlation coefficient = 0.59).

The MMAS-8 consists of eight items addressing various aspects of medication-taking behavior. Seven items utilize dichotomous yes/no response options, while one item employs a 5-point Likert scale. For scoring purposes, responses were transformed as follows: six items were coded as 1 for “yes” responses and 0 for “no” responses; one reverse-scored item was coded as 0 for “yes” and 1 for “no”; and the final item was scored using a graduated scale where responses of “never/rarely” = 1, “once in a while” = 0.75, “sometimes” = 0.5, “usually” = 0.25, and “always” = 0. The total MMAS-8 score ranges from 0 to 8, with higher scores indicating better medication adherence. Based on established cutoff points, scores were categorized as high adherence (8 points), medium adherence (6 to <8 points), and low adherence (<6 points).

The main exposure in this study was the therapeutic use of functional foods. We assessed the main exposure using a survey item that asked participants “Do you use supplements or Foods for Specified Health Uses (FOSHU) other than therapeutic drugs to lower blood pressure?” Based on their responses, participants were classified into two groups: those who reported using functional foods with the intention of treating hypertension (user group), and those who did not use such products for therapeutic purposes (non-user group). The survey question did not specify particular regulatory categories of functional foods. Participants determined whether a product qualified as a functional food based on their own understanding.

Based on previous systematic reviews on medication adherence in hypertensive patients^13^, covariates were selected as potential confounding variables and grouped into three categories: demographic factors (age, sex), socioeconomic factors (education, employment status, insurance type), and health-related factors (number of comorbidities, polypharmacy status, self-rated physical health). Variable definitions and coding procedures are detailed in the Supplementary table 1.

### Statistical analysis

Descriptive statistics were calculated to characterize the study population, with continuous variables presented as means and standard deviations, and categorical variables as frequencies and percentages. The distribution of MMAS-8 scores was examined overall and stratified by functional food use status. Scores were categorized as high adherence (8 points), medium adherence (6 to <8 points), and low adherence (<6 points).

To adjust for potential selection bias inherent in Internet-based survey data, all analyses applied inverse probability weighting (IPW). Weights were calculated as the inverse of the propensity score estimated via logistic regression, using covariates identical to those described in studies based on JACSIS 2020^14,15^ stratified by gender and age groups (15-19, 20-29, …, 70-79 years). For participants aged 20-79 years, covariates included socio-economic characteristics (residence area, marital status, education level, and home-ownership) and health-related characteristics (self-rated health and smoking status). For participants aged 15-19 years, covariates included socio-economic characteristics (residence area, education level, and home-ownership) and self-rated health, as marital status and smoking status were not applicable or available for this age group.

The primary analysis estimated risk ratios (RRs) for low adherence (MMAS-8 <6 points) associated with functional food use, using modified Poisson regression with a log link and robust standard errors. We fitted a sequence of multivariable models: Model 1 included the primary exposure (functional food use) only; Model 2 additionally adjusted for demographic factors (age and sex); Model 3 further incorporated socioeconomic factors (education level, employment status, insurance type, household income, and financial assets); and Model 4 was the fully adjusted model, including all demographic, socioeconomic, and health-related factors (comorbidity count, polypharmacy status, self-rated physical health), as well as region and prespecified hypertension self-management proxies. Weighted generalized linear models were used to combine the weighting with robust variance estimation.

We conducted subgroup analyses to examine potential effect modification across five key factors: smoking status, household income category, number of comorbidities, anxiety about blood pressure medication start/intensification, and experience of medication side effects. Interaction terms between functional food use and each subgroup variable were tested using the Wald test in the fully adjusted model (Model 4)Sensitivity analyses included (i) unweighted versions of the modified Poisson models and (ii) linear regression treating MMAS-8 as a continuous outcome to assess consistency of direction and magnitude.

All statistical analyses were conducted in R version 4.4.2 using the survey (version 4.4.2), sandwich (version 3.1.1), and lmtest (version 0.9-40) packages. Results were reported with 95% confidence intervals (CIs).

## Results

A total of 4,063 participants were included in the final analysis, comprising 586 (14.4%) functional food users and 3,477 (85.6%) non-users. The baseline characteristics of participants are presented in Table 1.

**Table 1.**
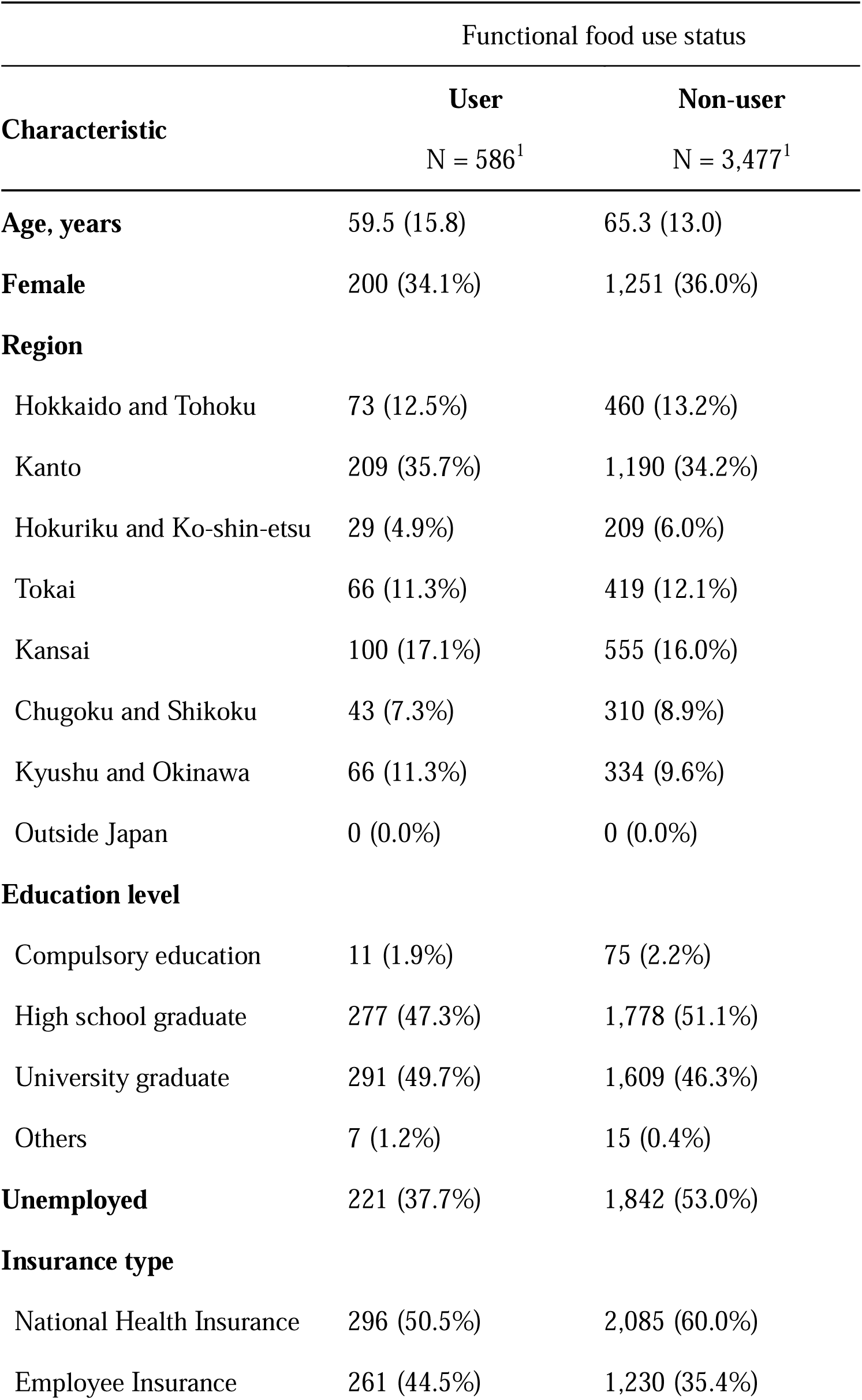
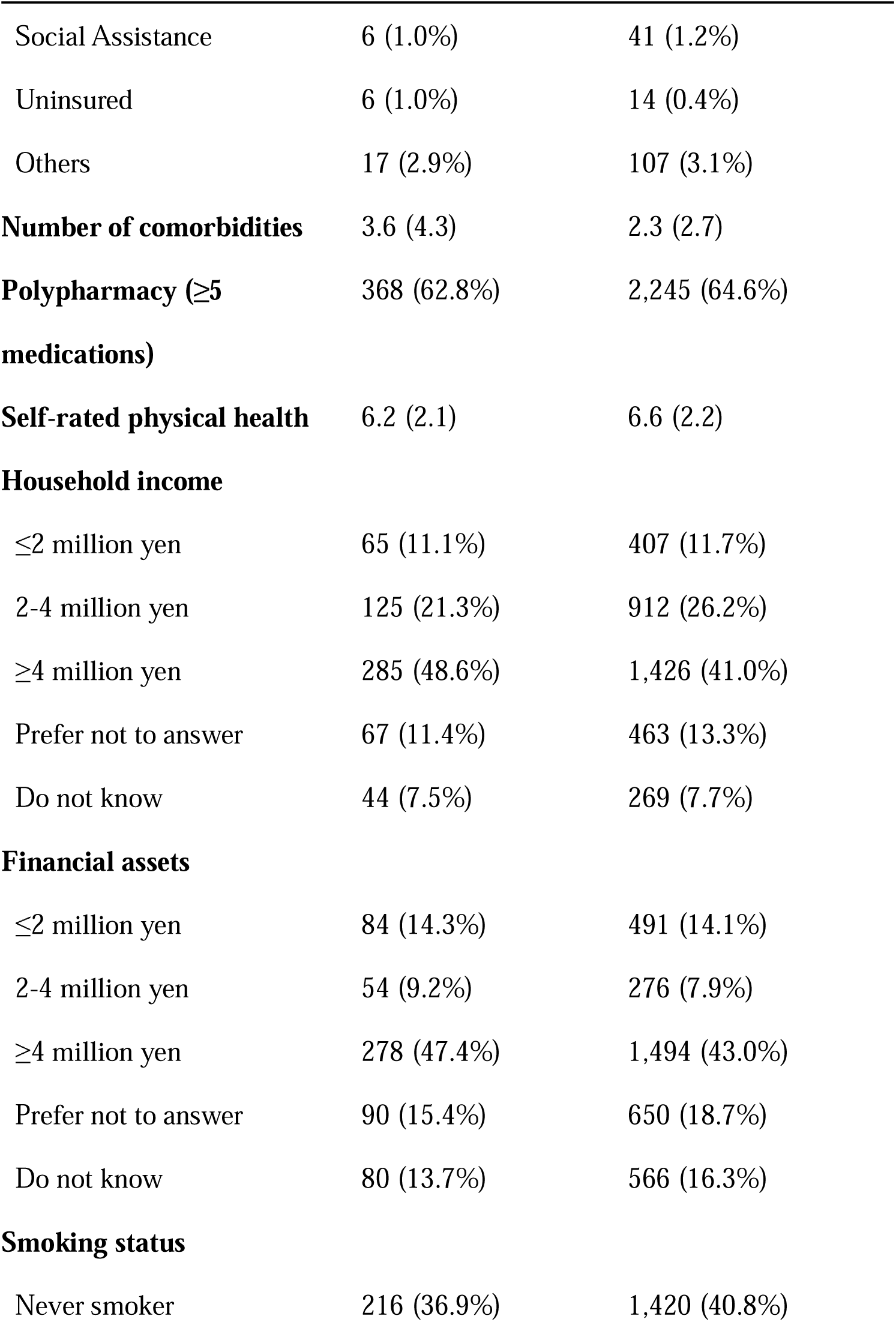
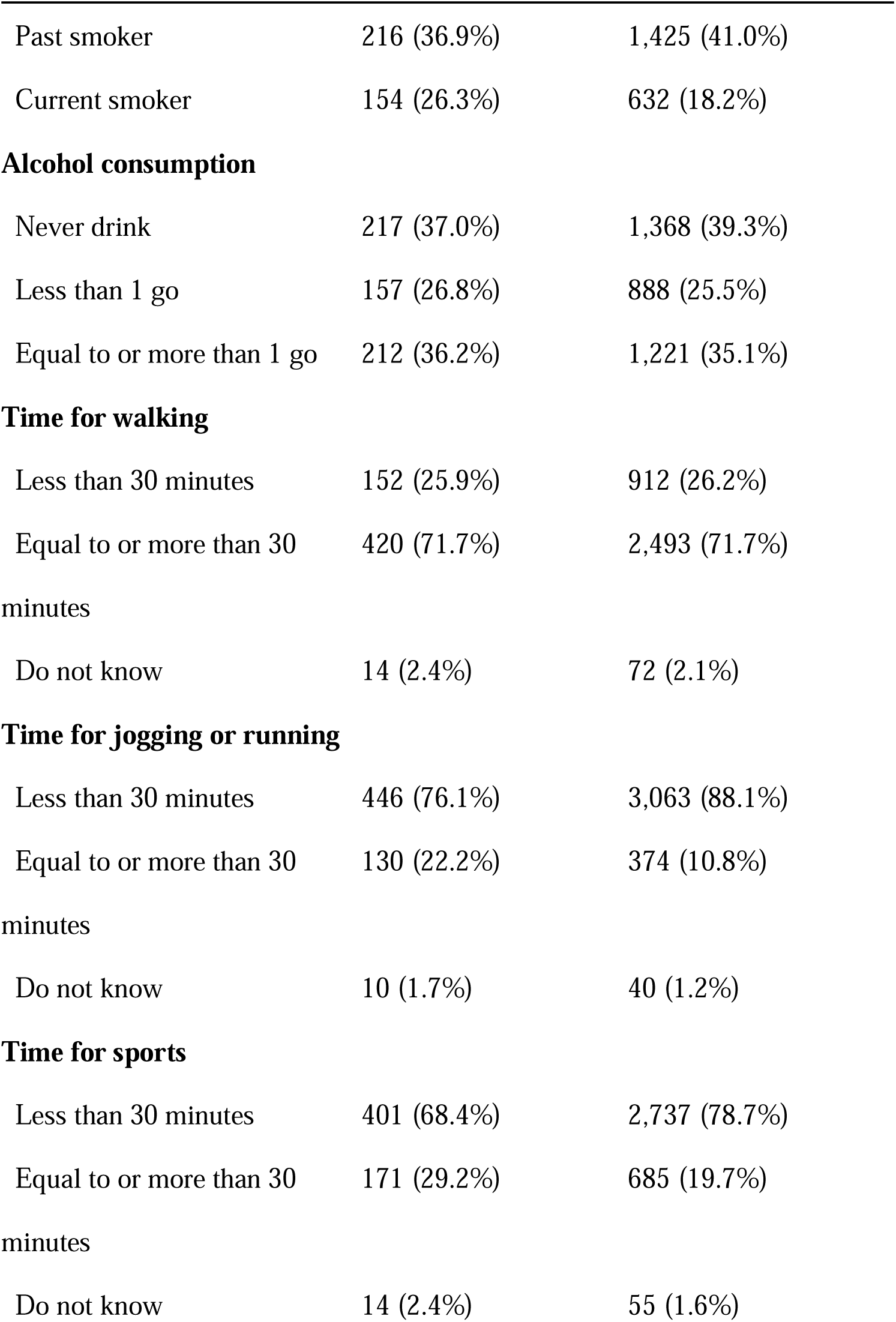
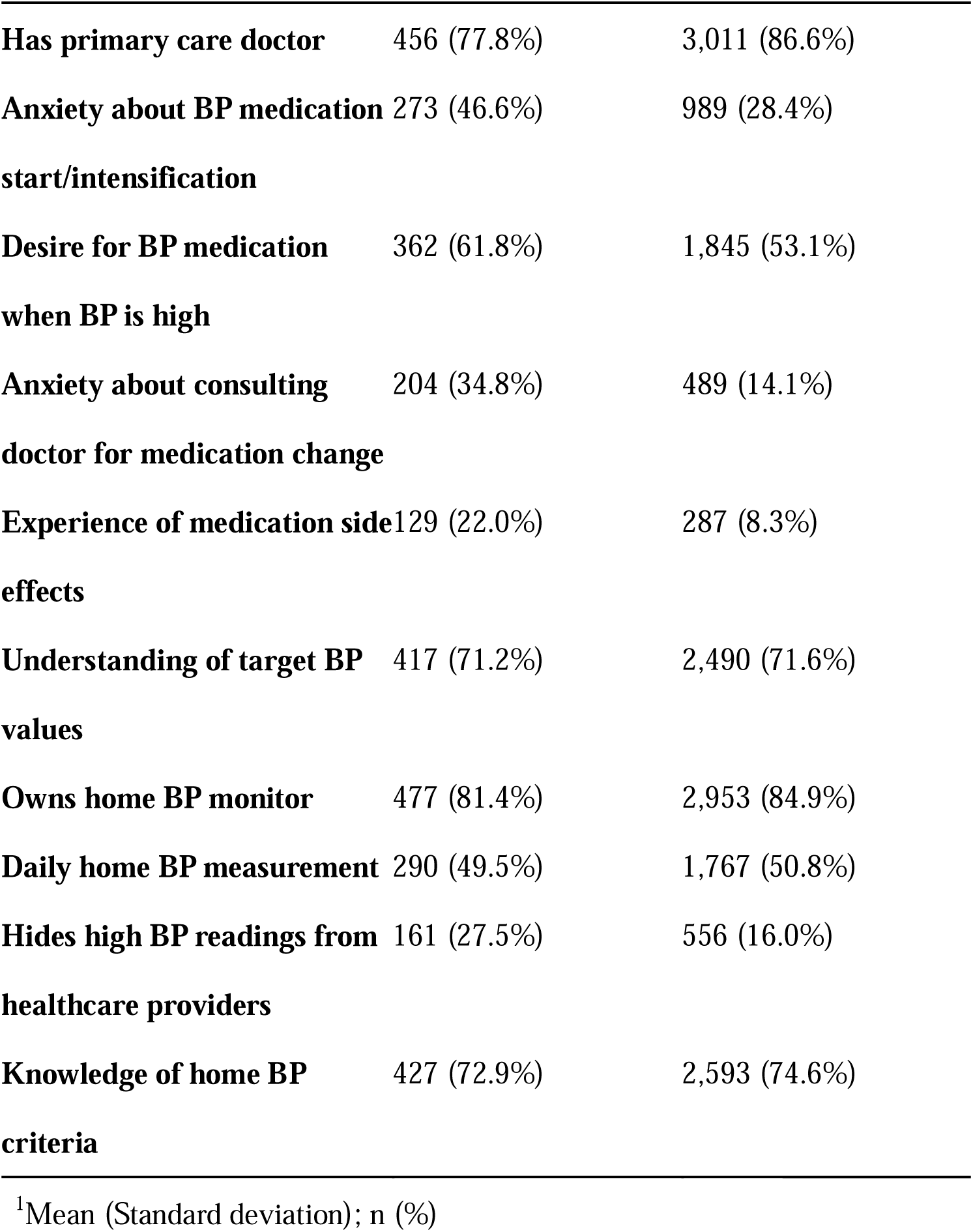
Baseline characteristics by functional food use status.

As for demographic differences, functional food users were younger on average (59.5 ± 15.8 years vs. 65.3 ± 13.0 years) and more likely to be employed (62.3% vs. 47.0%). The distribution of sex and geographic regions was relatively similar between groups. Regarding socioeconomic characteristics, functional food users demonstrated higher socioeconomic status, with a greater proportion having household income ≥4 million yen (48.6% vs. 41.0%) and financial assets ≥4 million yen (47.4% vs. 43.0%). They were also more likely to be current smokers (26.3% vs. 18.2%) and had a higher prevalence of cardiovascular comorbidities, including angina or myocardial infarction (15.7% vs. 8.2%) and stroke (11.9% vs. 5.4%) (Supplementary table 2).

Healthcare engagement patterns differed between groups. Functional food users were less likely to have a primary care doctor (77.8% vs. 86.6%) but showed greater anxiety about blood pressure medication initiation or intensification (46.6% vs. 28.4%) and consulting doctors for medication changes (34.8% vs. 14.1%). They also reported higher rates of medication side effect experiences (22.0% vs. 8.3%) and were more likely to hide high blood pressure readings from healthcare providers (27.5% vs. 16.0%).

The mean total MMAS-8 score was higher among functional food non-users compared to users (6.5 ± 1.7 vs. 5.7 ± 2.0, respectively), indicating better medication adherence in the non-user group. The standardized mean difference was 0.41. Individual item analysis revealed that functional food users demonstrated poorer adherence behaviors across most domains (Table 2). Notably, functional food users were more likely to cut back or stop medication without telling their doctor (22.7% vs. 11.2%) and to stop medication when feeling their blood pressure was under control (35.7% vs. 19.9%). The weighted data are shown in Supplementary Table 3.

**Table 2.** Medication Adherence Scale Responses by Functional Food User Classification. For items 1–7, numbers and percentages represent participants who answered ‘Yes’ to each question. For item 8, numbers and percentages represent the distribution across all response frequency categories (Never/rarely, Once in a while, Sometimes, Usually, All the time).

| Item | Functional food user | Functional food non-user |
| --- | --- | --- |
|  | N = 586<sup>1</sup> | N = 3477<sup>1</sup> |
| **Total score** | 5.7 (2.0) | 6.5 (1.7) |
| **Adherence Category** |  |  |
| **High ( = 8)** | 143 (24.4%) | 1,269 (36.5%) |
| **Middle (6–8)** | 180 (30.7%) | 1,254 (36.1%) |
| **Low (< 6)** | 263 (44.9%) | 954 (27.4%) |
| **Individual item** |  |  |
| **1. Do you sometimes forget to take your antihypertensive medication?** | 244 (41.6%) | 1,171 (33.7%) |
| **2. Over the past 2 weeks, were there any days when you did not take your antihypertensive medication?** | 185 (31.6%) | 689 (19.8%) |
| **3. Have you ever cut back or stopped taking your antihypertensive medication without telling your doctor because you felt worse when you took it?** | 133 (22.7%) | 388 (11.2%) |
|  | N = 586<sup>1</sup> | N = 3477<sup>1</sup> |
| 4. When you travel or leave home, do you sometimes forget to bring your antihypertensive medication? | 165 (28.2%) | 666 (19.2%) |
| 5. Did you take your antihypertensive medicine yesterday? | 502 (85.7%) | 3,161 (90.9%) |
| 6. When you feel like your blood pressure are under control, do you sometimes stop taking your antihypertensive medication? | 209 (35.7%) | 692 (19.9%) |
| 7. Do you ever feel hassled about sticking to your treatment plan? | 256 (43.7%) | 1,076 (30.9%) |
| 8. How often do you have difficulty remembering to take all your antihypertensive medication? |  |  |
| All the time | 6 (1.0%) | 19 (0.5%) |
| Usually | 7 (1.2%) | 19 (0.5%) |
| Sometimes | 32 (5.5%) | 106 (3.0%) |
|  | N = 586<sup>1</sup> | N = 3477<sup>1</sup> |
| Once in a while | 154 (26.3%) | 619 (17.8%) |
| Never/rarely | 387 (66.0%) | 2,714 (78.1%) |

<sup>1</sup> Mean (Standard deviation); n (%)

In weighted modified Poisson models, adjusting for demographic, socioeconomic, and health-related factors, functional food use was associated with a higher risk of low adherence: RR 1.24 (95% CI, 1.04–1.48; Table 3).

**Table 3.** Association between functional food use and low medication adherence (Morisky medical adherence scale-8 < 6): weighted modified Poisson regression.

| **Model** | **Risk ratio (95% confidence interval)** |
| --- | --- |
| Model 1 | 1.44 (1.22–1.71) |
| Model 2 | 1.41 (1.19–1.67) |
| Model 3 | 1.35 (1.13–1.62) |
| Model 4 | 1.24 (1.04–1.48) |

Note:
- Risk ratios $> 1$ indicate a higher risk of low adherence among functional food users compared with non-users.

The subgroup analyses revealed no substantial differences in the association between functional food use and medication adherence across the examined factors (Supplementary Table 4). The risk ratios remained generally consistent across subgroups defined by smoking status, household income category, number of comorbidities, anxiety about blood pressure medication start/intensification, and experience of medication side effects (all p-values for interaction >0.05).

As a sensitivity analysis, an unweighted modified Poisson model, adjusting for demographic, socioeconomic, and health-related factors, yielded RR 1.18 (95% CI, 1.06–1.32; Supplementary Table 5). In an additional analysis treating MMAS-8 as a continuous outcome with the same covariate adjustment, the adjusted mean difference (users minus non-users) was −0.30 points (95% CI, −0.44 to −0.16, Supplementary Table 6).

## Discussion

This study examined the relationship between functional food use with therapeutic intent and medication adherence among 4,063 hypertensive patients in Japan (586 users; 3,477 non-users). In weighted, fully adjusted modified Poisson models, functional food use was associated with a higher risk of low adherence (MMAS-8 < 6): RR 1.24 (95% CI, 1.04–1.48). Unweighted models produced similar estimates (RR 1.18; 95% CI, 1.06–1.32). When MMAS-8 was modeled as a continuous outcome, the adjusted mean difference (users minus non-users) was −0.30 points (95% CI, −0.44 to −0.16), consistent with the primary analysis..

Our study suggests that, among patients with treated hypertension, functional food use is associated with low medication adherence. Previous studies investigating the relationship between dietary supplement use and medication adherence have generally reported no association between these factors^7,8^. However, these earlier investigations examined general dietary supplement use rather than products specifically marketed or perceived as having blood pressure-lowering effects. Our study focused on functional foods that participants believed could help treat their hypertension, representing a fundamentally different therapeutic intent compared to general supplementation. This distinction in purpose may explain the divergent findings from previous research..

The reduced medication adherence observed among functional food users may be attributed to underlying distrust or anxiety toward medications and healthcare providers. Although both groups showed similar ratios of having a primary care doctor (77.8% vs 86.6%), functional food users demonstrated markedly different attitudes toward medical treatment. They consistently reported higher ratios of anxiety toward medications and healthcare consultations, greater experiences of medication side effects, and more frequent concealment of elevated blood pressure readings from healthcare providers. These findings suggest that the use of functional foods may represent a coping mechanism for patients who harbor reservations about conventional pharmacological approaches, potentially leading them to view these products as preferable alternatives to prescribed medications rather than complementary interventions. While some patients may turn to functional foods due to adverse effects from specific medications, it is crucial to recognize that experiencing side effects with one medication does not preclude the safe use of alternative antihypertensive agents. Rather than resorting to inappropriate functional food use, patients should engage in discussions with healthcare providers to explore alternative medications or dosing regimens that may be better tolerated while maintaining therapeutic efficacy.

Two approaches may be considered to reduce the inappropriate use of functional foods observed in our study. First, and most importantly, consumer-targeted educational interventions are warranted given the demographic characteristics of functional food users in our cohort. Compared to non-users, functional food users demonstrated higher socioeconomic status with greater income levels, were younger (mean age 59 vs 65 years), had more cardiovascular comorbidities, and exhibited higher smoking rates. These findings suggest that users may have lower health literacy despite their higher socioeconomic status. Moreover, they may be more susceptible to exaggerated or misleading health claims, indicating a need for targeted educational initiatives. Specifically, efforts should focus on promoting accurate understanding of functional foods among consumers. The Food Safety Commission of Japan has already developed 19 key messages regarding so-called health foods,^17^ including the critical statement that “health foods cannot replace medications, so you should not stop taking prescribed drugs” and that “no food products have been scientifically proven to extend healthy life expectancy.” Broader dissemination of these evidence-based messages could help correct misconceptions about the therapeutic role of functional foods. Second, consideration might also be given to reviewing current regulatory approaches to health food advertising. While existing regulations already prohibit claims that could lead consumers to mistake these products for pharmaceuticals, our findings suggest that misunderstandings may persist among consumers. Potential measures could include strengthening mandatory labeling requirements to more explicitly state “not for use as disease treatment” or similar warnings, though further research would be needed to evaluate the effectiveness of such approaches.

Several limitations should be acknowledged in interpreting our findings. First, we were unable to adjust for hypertension duration or disease severity, which may influence both functional food use and medication adherence patterns. However, our analysis did include comorbidities such as stroke and cardiovascular disease history, providing indirect adjustment for disease severity. Second, our study was limited to treated hypertensive patients currently receiving medication. The impact of functional food use on unscreened individuals or those with diagnosed but untreated hypertension requires further investigation. Third, we focused specifically on functional foods used with the intention of managing blood pressure rather than evaluating general functional food consumption. Additionally, our survey questionnaire referred to “FOSHU (Foods for Specified Health Uses) and supplements” without distinguishing between specific categories of functional foods, and we could not determine whether the reported products fell within a particular regulatory framework (e.g., FOSHU, Foods with Function Claims) or were unregulated health foods. Therefore, our findings do not provide insights into the broader role or value of functional foods in health maintenance, limiting our ability to discuss the overall merits of functional food use. Fourth, we did not collect direct blood pressure measurements or control status. While this limits inferences about clinical endpoints, our primary outcome—self-reported medication adherence—captures a proximal behavioral domain that influences blood pressure management. Therefore, the observed association between functional food use and adherence remains interpretable as a behavioral finding and does not alter our main conclusions. Lastly, while we assessed participants’ understanding of having target blood pressure values, we did not evaluate their knowledge of specific target values.

## Conclusion

This cross-sectional study found an association between functional food use with therapeutic intent and higher risk of low medication adherence among patients with treated hypertension. The observed association appears to reflect underlying anxiety and distrust toward conventional medical treatments. While these findings suggest a concerning pattern where some patients may substitute evidence-based antihypertensive therapy with functional foods, the cross-sectional nature of our study limits our ability to establish causality. Nevertheless, these observations may warrant consideration of targeted interventions. Educational approaches to improve health literacy regarding the appropriate role of functional foods could be beneficial, alongside potential review of current health food advertising and labeling practices to ensure consumers receive clear, evidence-based information. Future longitudinal research should examine the temporal relationship between functional food use and medication adherence, investigate the impact on clinical outcomes such as blood pressure control, and explore interventions to optimize both functional food use and medication adherence in hypertensive patients.

## Data Availability

Access to the data is governed by the study investigators and requires permission. Researchers interested in using the data should submit a research proposal that is consistent with ethical approval, confidentiality agreements, and data management protocols. To apply for data access or to obtain more information about the study, please contact the following researcher: Dr. Takahiro Tabuchi, MD, PhD Dr. Tabuchi can provide further details about the application process, available data, and potential collaborations.

## Acknowledgements

We thank Donald E. Morisky, ScD, ScM, MSPH, for granting permission to use the MMAS-8 scale under a formal license agreement.

## References

1 NCD Risk Factor Collaboration (NCD-RisC). Worldwide trends in hypertension prevalence and progress in treatment and control from 1990 to 2019: a pooled analysis of 1201 population-representative studies with 104 million participants. Lancet 2021; 398: 957–980.

2 Hamdidouche I, Jullien V, Boutouyrie P, Billaud E, Azizi M, Laurent S. Drug adherence in hypertension: from methodological issues to cardiovascular outcomes. J Hypertens 2017; 35: 1133–1144.

3 Arai S. Studies on functional foods in Japan--state of the art. Biosci Biotechnol Biochem 1996; 60: 9–15.

4 Henry CJ. Functional foods. Eur J Clin Nutr 2010; 64: 657–659.

5 Pandey D. Functional Food Ingredients Market Size, Share, and Trends 2025 to 2034. Precedence research. https://www.precedenceresearch.com/functional-food-ingredients-market. Accessed May 21, 2025

6 Chiba T. Patients Are Using Dietary Supplement for the Treatment of Their Diseases without Consultation with Their Physicians and Pharmacists. Pharmacy (Basel) 2023; 11. doi:10.3390/pharmacy11060179

7 Eussen SRBM, Verhagen H, Klungel OH, Garssen J, van Loveren H, van Kranen HJ, Rompelberg CJM. Functional foods and dietary supplements: products at the interface between pharma and nutrition. Eur J Pharmacol 2011; 668 Suppl 1: S2–9.

8 Fadzil SK, Omar M, Tohit N. Practice of dietary supplements and its influence towards treatment adherence among chronic disease patients. International journal of public health research 2018; 8: 998–1005.

9 Tabuchi T, Shinozaki T, Kunugita N, Nakamura M, Tsuji I. Study Profile: The Japan “Society and New Tobacco” Internet Survey (JASTIS): A Longitudinal Internet Cohort Study of Heat-Not-Burn Tobacco Products, Electronic Cigarettes, and Conventional Tobacco Products in Japan. J Epidemiol 2019; 29: 444–450.

10 Tousen Y, Kondo T, Chiba T, Ishimi Y. Regulation of the Food Labelling Systems for Health and Nutrition in Japan and Associated Role of the National Institute of Health and Nutrition. Jpn J Nutr Diet 2020; 78: S80–S90.

11 Tanaka M, Kawakami A, Maeda S, Kunisaki R, Morisky DE. Validity and reliability of the Japanese version of the Morisky Medication Adherence Scale-8 in patients with ulcerative colitis. Gastroenterol Nurs 2021; 44: 31–38.

12 Muntner P, Joyce C, Holt E, He J, Morisky D, Webber LS, Krousel-Wood M. Defining the minimal detectable change in scores on the eight-item Morisky Medication Adherence Scale. Ann Pharmacother 2011; 45: 569–575.

13 Uchmanowicz B, Jankowska EA, Uchmanowicz I, Morisky DE. Self-reported medication adherence measured with Morisky medication adherence scales and its determinants in hypertensive patients aged ≥60 years: A systematic review and meta-analysis. Front Pharmacol 2019; 10: 168.

14 Miyawaki A, Tabuchi T, Tomata Y, Tsugawa Y. Association between participation in the government subsidy programme for domestic travel and symptoms indicative of COVID-19 infection in Japan: cross-sectional study. BMJ Open 2021; 11: e049069.

15 Koyama T, Takeuchi K, Tamada Y, Aida J, Koyama S, Matsuyama Y, Tabuchi T. Prolonged sedentary time under the state of emergency during the first wave of coronavirus disease 2019: Assessing the impact of work environment in Japan. J Occup Health 2021; 63: e12260.

16 Mirniam A-A, Habibi Z, Khosravi A, Sadeghi M, Eghbali-Babadi M. A clinical trial on the effect of a multifaceted intervention on blood pressure control and medication adherence in patients with uncontrolled hypertension. ARYA Atheroscler 2019; 15: 267–274.

17 The Food Safety Commission of Japan (FSCJ). Iwayuru “Kenkou Shokuhin” ni Kansuru Message. 2015. https://www.fsc.go.jp/osirase/kenkosyokuhin.data/kenkosyokuhin_message.pdf. Accessed July 29, 2025

